# Survey of Direct and Indirect Effects of COVID-19 on Eyes and the Common Ocular Manifestations

**DOI:** 10.1101/2021.10.18.21265130

**Authors:** Dianeh Rabi, Razan Rabi, Arkan Jarrar, Sarah Mharma, Aya Jaradat, Shatha Bzoor

**Author notes:** ***Correspondence* Razan rabi**, MD, Intern, Rafidia Hospital, Ministry of Health., *Email*, *Mob 00 970 592055585 *, Adress Rafidia, Nablus, Palestine. **Author contributions**. These authors also contributed equally to this work (Dianeh Rabi, Razan Rabi). These authors also contributed equally to this work (Sarah Mharma, Aya Jaradat, Shatha Bzoor).

## Abstract

**Purpose:** Ocular manifestations were reported in many recent observations that studied either the effect of COVID-19 directly on eyes or of face mask use. Hence, this study aimed to investigate the effect of COVID-19 on the eyes and make a clear comparison of its direct and indirect effect from face mask-wearing.

**Methods:** This was a cross-sectional study of both written and web-based questionnaires, distributed among a group of COVID-19 patients and a matched control group, the questionnaire consisted of common demographic data, COVID-19 infection history and its symptoms, focusing on ocular symptoms and the presence of conditions related to or cause eye symptoms. As well as the use of face masks that were assessed in terms of the complained ocular manifestation

**Results:** Of 618 participants, 252 had COVID-19 and 366 never had COVID-19. Ocular manifestation among COVID-19 incidence was 44%, significantly higher than non-infected participants’ incidence (35.8%), adjusted odds ratio, 95% confidence interval (AOR, 95%CI); 1.45 (1.02-2.06)). Eye discharges (p-value = 0.033) and photosensitivity (p-value = 0.003) were noted more commonly among COVID-19 participants compared to healthy control. When comparing long periods of face mask use with each ocular symptom; dry eye based on OSDI, forging body sensation, eye pain and eye discharges, were found significantly common among extended periods of face mask use.

**Conclusion:** COVID-19 pandemic affected eyes, both directly from the virus or from its preventive measure of face mask use.

## Introduction

Coronavirus disease (COVID-19) is an infectious disease caused by the newly discovered coronavirus SARS-CoV-2. It is a non-segmented virus, which has a large single□stranded RNA virus genome, capped and polyadenylated.(1) It is one of the highly pathogenic β□coronaviruses which infect humans and cause a wide range of symptoms that can be fatal. Globally, at this time, there have been more than 151 million confirmed cases of COVID-19, including more than 3 million deaths.(2)

COVID-19 virus spreads primarily through droplets of saliva or discharges from the nose when an infected person coughs or sneezes. However, transmission by tears as an alternative mode of transmission had been reported by the finding of the virus on tears suggesting this possibility, but this remains unclear.(3,4) Xia et al. assessed 30 patients with COVID-19, only one tested sample of tears and conjunctival secretion from a patient with conjunctivitis had a positive reverse transcriptase-polymerase chain reaction (RT-PCR) results. (4) Many reports hypothesized that direct inoculation of the mucous membrane of the conjunctival surface at a site of infected droplets or through the nasolacrimal duct can act as a channel for viral migration.(5)

COVID-19 is primarily a respiratory illness, with the most common symptoms are respiratory ones, but few reports found that some individuals with the disease had presented with ocular manifestations, (6–10) though the exact pathogenic mechanisms of ocular infection were still unknown. A study in China showed that among children diagnosed with the virus, ocular manifestations were the initial symptoms in 22.7%, with the most common ocular complaints of these were eye rubbing, conjunctival discharge, and conjunctival congestion. (11) On the other side, observations on the effect of face masks wearing revealed that mask-associated ocular manifestations such as dryness and irritation prove to be prevalent among all mask wearers, and particularly in long periods of use. As a result, it suggested the use of lubricant eye drops and masks for eye protection. (12,13)

Early diagnosis of COVID-19 is the most critical step to treat the infection. This necessitates understanding all its signs and symptoms, which frame the prompt diagnosis. This study had aimed to investigate the frequency of ocular manifestations and delineate the direct effect of COVID-19, in which the virus can either infect the eyes directly or from the indirect effects of its preventive measures namely through the face mask use, that can cause eye dryness and irritation. Assessing this association in regards to other non-COVID-19 related factors that affect the eyes.

## Material and methods

### Study design and sampling

This was a cross-sectional survey, that was distributed during May 2021 by both written and a web-based questionnaire through various Palestinian individual or public media platforms, and the written ones were distributed in a primary and a secondary health care setting in Palestine to target as many COVID-19 participants and matched controls, as well to prevent bias of web-based design by involving more older and illiterate groups. The study included COVID-19 currently infected or recovered participants, confirmed with nasopharyngeal swab samples for SARS-CoV-2. Along with another control group that never had or was suspected to have COVID-19, matched for age and gender. Targeting a sample size of more than 226 for COVID-19 patients with a case: control ratio of 1:2, considering a 95% confidence interval and 90% power with an expected frequency of 18% vs 9% among COVID-19, general participants respectively. (6,14,15) Participants who were older than 18 years and had COVID-19 within 6 months were recruited with a matched healthy control group. Excluding those who had chronic eye diseases that are known to cause dryness or inflammatory manifestations like chronic conjunctivitis, and those reported having chronic ocular complaints of more than 6 months (for a purpose of recall bias prevention, the study period was for the past 6 months), as well as those who had strong susception of COVID-19 but no confirmatory tests. The study recruited 715 (302 COVID-19) participants, after the exclusion of 97 participants, the total number was 618 (252 COVID-19).

Before beginning the questionnaire, a request to declare voluntary consent was provided stating voluntary acceptance to participate, after describing the study and its objectives with ensuring the participants’ data privacy. The study was approved to be in accordance with the Declaration of Helsinki by AL-Najah National University’ Ethics Committee.

### Measures and variables

The questionnaire consisted of two divisions, one for currently or previously had COVID-19, and another for those who never had COVID-19, sharing the following sections; demographics and personal data included; age, gender, address, education, and employment status, medical history, and smoking state. Protection measurements, particularly face mask practices, and specific questions regarding eye findings, including the main question of whether had any defined ocular complaints within the past 6 months (eye irritation symptoms; burning, itching, discharge, dryness, blurry vision, forging body sensation, euphoria, seeing floater and double vision) and other eye-related data like using contact lenses, any previous chronic eye diseases (refractive errors, cataract, chronic conjunctivitis, etc.) and if any used eye-drops. Known factors affecting eyes were assessed as well, like duration of electronic device use (smartphone, laptop, etc.), duration of short-distance reading, and long office hours. Eye dryness was assessed particularly, on all participants using the Ocular Surface Disease Index (OSDI) questionnaire within a separate section, after it was translated into the Arabic language, ensuring that the translated version of the OSDI was in accordance with its publisher Allergan Inc. (Irvine, CA, USA). Each division of the questionnaire had asked questions accordingly; for COVID-19 patients, it had questions regarding the time of COVID-19 diagnosis, the complained COVID-19 symptoms, infection confirmation tests, severity of illness, and the needed management. Moreover, the timing of eye manifestations in relation to the starting day since COVID-19 beginning or recovery was questioned. While non-COVID-19 participants, general COVID-19 symptoms history, to assure not to be suspected of having COVID-19, were addressed in a side with the other shared items.

### Definitions

Each participant was included in the COVID-19 group if either confirmed by RT-PCR nasopharyngeal swab or rapid antigen test. The severity of COVID-19 was classified as asymptomatic, mild (did not need any therapy besides in-need painkillers or vitamins), moderate (needed home therapy), severe (needed hospitals), and critical (entered intensive care unit (ICU)). The ocular surface disease index (OSDI) is a questionnaire to determine the severity of eye dryness from 12 items, in which a score > 12 is defined as dryness, with values 12-22, 23-32, and 32 > is mild, moderate and severe dryness respectively.(16) The term indirect effect of COVID-19 was given for the need for face mask use, and its duration of use was sorted as short duration (less than 4 hours), moderate (4-6 hours), and long period (more than 6 hours).

### Statistical Analysis

All variables were displayed as frequencies or mean ± standard deviation (SD). Ocular manifestations were compared between COVID-19 and non-COVID-19 infected participants as well as between face mask periods of use. Investigating the association of these main factors independently from each other, and after adjusting for possible confounders, those were; hours of short-distance reading duration, electronic device usage, long office hours, contact lenses use, and existing eye diseases. Confounders were tested and adjusted for, by using binary logistic regressions. Moreover, subgroup analysis was performed comparing COVID-19 participants with and without ocular findings, in relation with the other study variables, by chi-square, Exact fisher’s test and independent t-test as required. P-value was set at 0.05.

## Results

Of the total 618 recruited participants, 252 had COVID-19 and 366 never had COVID. in which 57% of the COVID-19 group were aged between 18-30 years, 59% were females, 13% had chronic diseases, 25% were smokers and 16% had chronic eye diseases (Table 1). Among COVID-19 infected participants, 9.5% of them were currently infected, while 46.5% and 53.1% were infected during the past 3 months, and before 3-6 months respectively. 55.6% of COVID-19 patients had confirmation of diagnosis by RT-PCR and the remaining were confirmed by the rapid test. Most cases (85%) of COVID-19 in the study had a mild-moderate illness, whereas severely infected cases accounted for 3.2% and only one case was critical (received ICU). The most reported non-ocular COVID-19 symptom was headache (62%). Other common ones were; arthralgia (32%), myalgia (31%), fever (19.8%), and chills (22%).

**Table 1.** Baseline characteristics of COVID-19 participants and non-COVID-19 controls.

| **Characteristics** | **Total**<br>**(n=618),**<br>**n (%)** | **COVID-19**<br>**(n=252), n (%)** | **Non-COVID-19**<br>**(n=366), n (%)** |
| --- | --- | --- | --- |
| **Age, year** |  |  |  |
| 18-30 | 326 (52.8%) | 144 (57.4%) | 182 (49.7%) |
| 31-45 | 160 (25.9%) | 62 (24.7%) | 98 (26.8%) |
| 46-65 | 103 (16.7%) | 32 (12.7%) | 71 (19.4%) |
| >65 | 28 (4.5%) | 13 (5.2%) | 15 (4.1%) |
| **Gender** |  |  |  |
| Female | 368 (59.5%) | 155 (61.5%) | 213 (58.2%) |
| **Educational level** |  |  |  |
| Higher educations | 46 (7.5%) | 18 (7.1%) | 28 (7.7%) |
| Bachelor's degree | 391 (63.4%) | 172 (68.3%) | 219 (60%) |
| High school | 127 (20.6%) | 45 (17.9%) | 82 (22.5%) |
| Less than high school | 49 (7.9%) | 15 (6%) | 34 (9.3%) |
| Non-literate | 4 (0.6%) | 2 (0.5%) | 2 (0.5%) |
| **Employment status** |  |  |  |
| Employed | 232 (37.5%) | 98 (38.9%) | 134 (36.6%) |
| Students | 181 (29.3%) | 91 (32.9%) | 98 (26.8%) |
| Unemployed | 194 (31.4%) | 67 (26.6%) | 127 (34.7%) |
| Retired | 11 (1.8%) | 4 (1.6%) | 7 (1.9%) |
| **Health care worker** | 151 (24.4%) | 66 (26.2%) | 85 (23.2%) |
| **Long office hours** | 178 (8.8%) | 86 (34.1%) | 92 (25.1%) |
| **Chronic diseases** | 99 (16%) | 32 (12.7%) | 67 (18.3%) |
| Hypertension | 45 (7.3%) | 12 (4.8%) | 33 (9%) |
| Diabetes | 57 (9.2%) | 22 (8.7%) | 35 (9.6%) |
| CVD | 25 (4%) | 13 (5.2%) | 12 (3.3%) |
| **Chronic eye diseases** | 106 (17.2%) | 41 (16.3%) | 65 (17.8%) |
| Diabetic retinopathy | 12 (1.9%) | 3 (1.2%) | 9 (2.5%) |
| Refractive errors | 87 (14.1%) | 35 (13.9%) | 52 (14.2%) |
| Cataract | 4 (0.6%) | 3 (1.5%) | 1 (0.2%) |
| **Contact lenses use** | 29 (4.7%) | 12 (4.8%) | 17 (4.6%) |
| **Used eye-drops** | 87 (14.1%) | 44 (17.5%) | 43 (11.7%) |
| **Smoking status** |  |  |  |
| Current smoker | 149 (24.1%) | 63 (25%) | 86 (23.5%) |
| Previous smoker | 13 (2.1%) | 6 (2.4%) | 7 (1.9%) |
| **Electronic devices usage** |  |  |  |
| **/day** | 132 (21.4%) | 53 (21%) | 79 (27.6%) |
| Less than 2 hours | 318 (51.5%) | 140 (55.6%) | 178 (48.8%) |
| 2-6 hours | 167 (27.1%) | 59 (23.4%) | 108 (29.6%) |
| More than 6 hours |  |  |  |
| **Hours of reading /day** |  |  |  |
| Less than 2 hours | 397 (64.3%) | 172 (68.3%) | 225 (61.6%) |
| 2-6 hours | 160 (25.9%) | 63 (25%) | 97 (26.6%) |
| More than 6 hours | 60 (9.7%) | 17 (6.7%) | 43 (11.8%) |
| **Hours of mask wearing/day** |  |  |  |
| None | 68 (11.2%) | 30 (12%) | 38 (10.6%) |
| 1-2 hours | 223 (36.6%) | 91 (36.4%) | 132 (36.8%) |
| 2-4 hours | 112 (18.4%) | 46 (18.4%) | 66 (18.4%) |
| 4-6 hours | 103 (16.9%) | 45 (18%) | 58 (16.2%) |
| > 6 hours | 103(16.9%) | 38 (15.2%) | 65 (18.1%) |
| CVD; cardiovascular diseases |  |  |  |

The effects of COVID-19 on eyes were demonstrated in Table 2. Ocular manifestations reported in higher frequencies among COVID-19 infected participants compared to never infected (44% vs 35.8%), adjusted odds ratio, 95% confidence interval (AOR, 95%CI); 1.45 (1.02-2.06)), exclusive symptoms that were significantly more common among COVID-19 compared to other were; eye discharge, photosensitivity, which were significantly more reported among COVID-19 participants p-value = 0.033 and 0.003 respectively, while eye dryness was significant when was questioned directly (p-value= 0.046) it was not based on OSDI score. (Table 2).

**Table 2.**
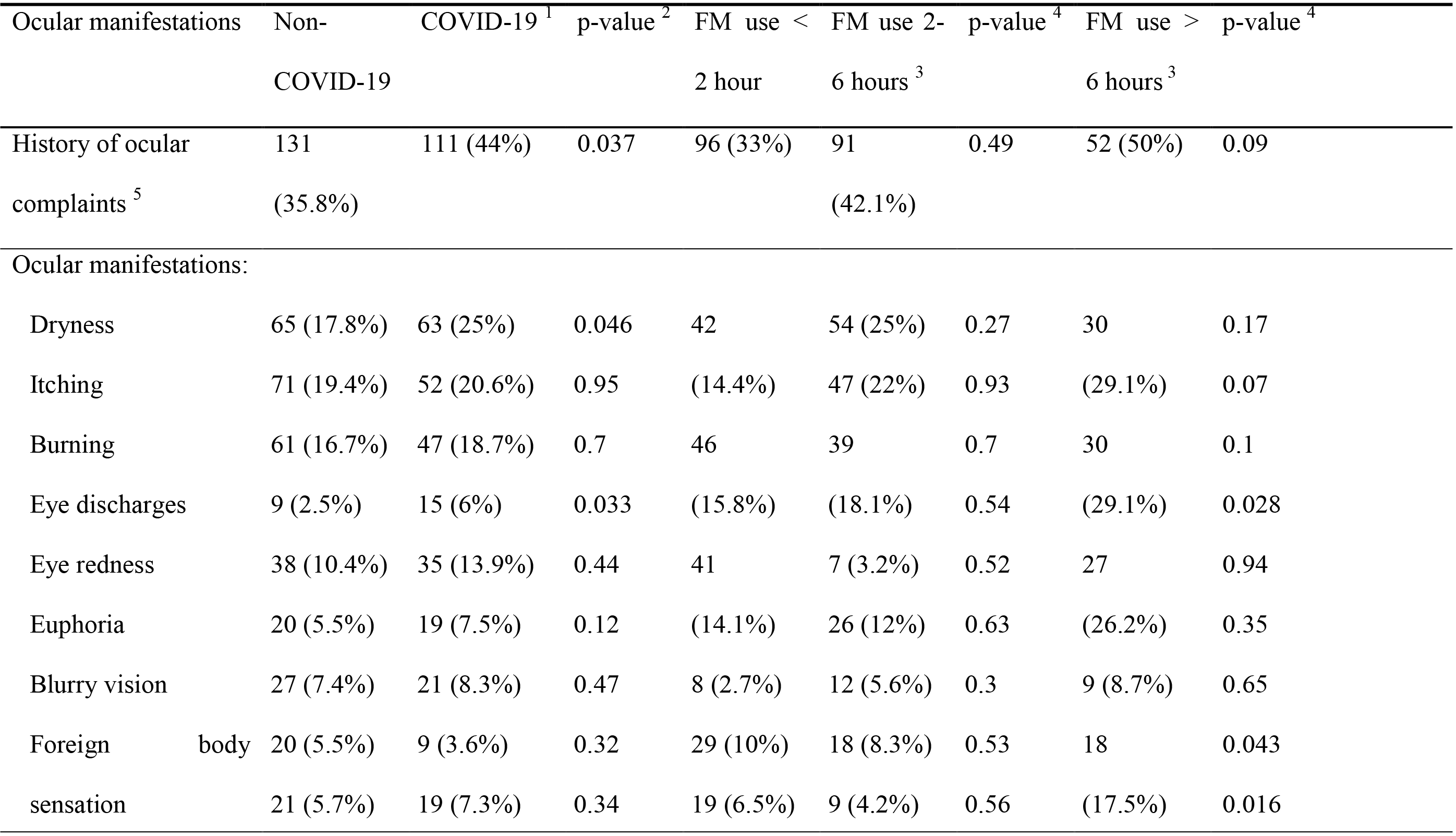
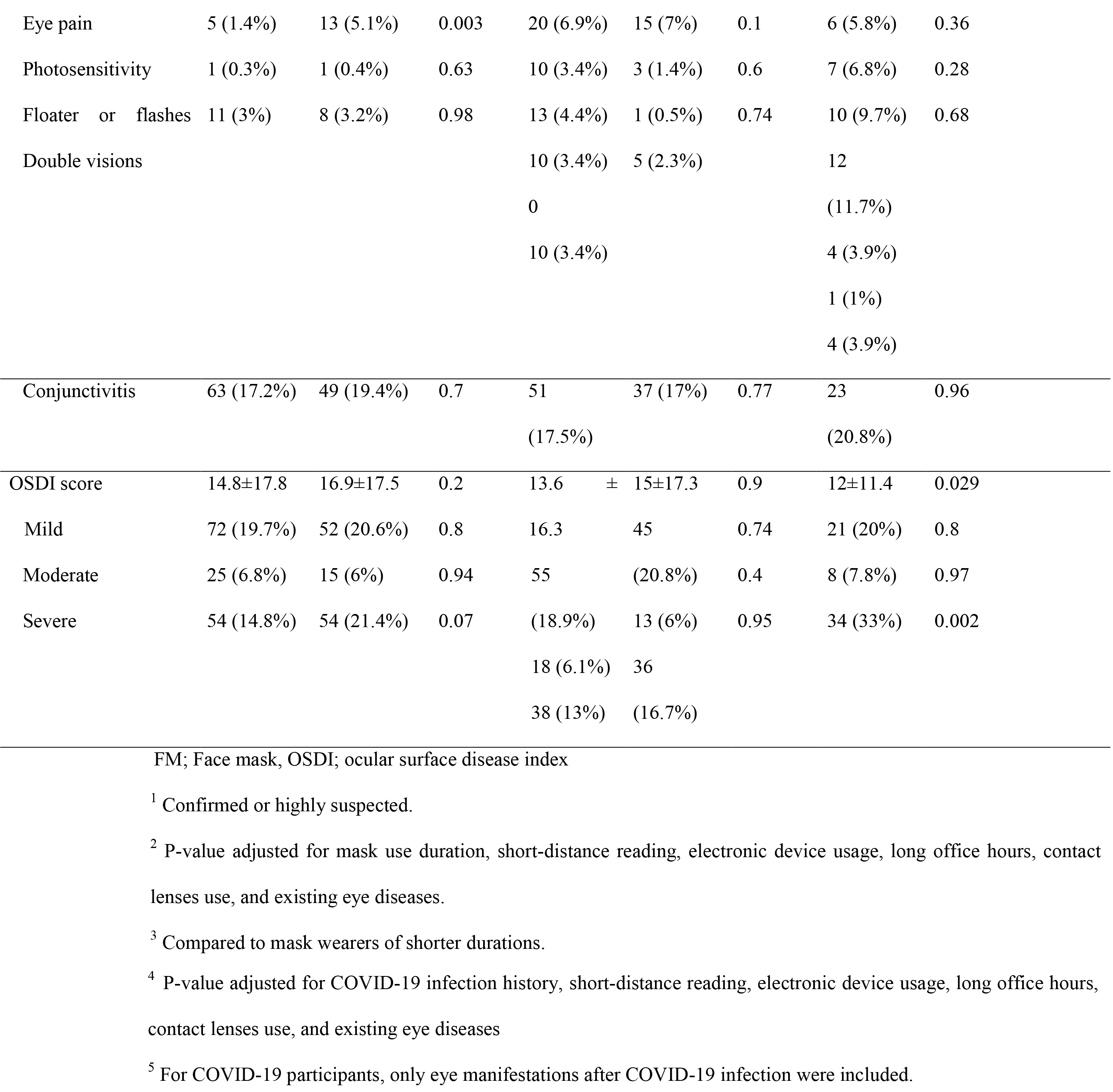
Ocular manifestations associated with COVID-19 directly and indirectly, by comparing between COVID-19 status and periods of face mask use. (n=618)

Furthermore, the indirect effect namely the face mask use, though the overall incidence of any ocular manifestations was significantly more reported among face mask wearers of extended periods before adjusting (p<0.001), significance was lost after adjusting for reading hours, smartphone usage hours, chronic eye diseases, contact lenses use and the COVID-19 state, in which for long and moderate face mask periods of use in comparing to short periods, adjusted p-value were 0.49 for moderate periods and 0.09 for long periods (50% vs 42.1% vs 33% for long, moderate, and short face mask use respectively). However, among individual signs or symptoms, a significantly higher frequency among long periods face mask use was reported for eye-pain (AOR, 95% CI; 2.5, (1.2-5.3)), eye-discharges (AOR, 95% CI; 2.8 (1.1-6.8)), forging body sensation (AOR, 95% CI; 2.4 (1.03-5.5) and eye dryness by OSDI (AOR, 95% CI; 1.8 (1.1-2.9)) (Table 2).

Of 252 COVID-19 patients, 88 had ocular manifestations after getting the infection, in which 40% complained of these manifestations during COVID-19 infection (32% of them remained after COVID-19 recovery). While 13.3% had these manifestations only after recovery from COVID-19. The remaining had reported having ocular manifestations before and after getting the disease with 8% of them having unfamiliar ocular complaints after it. The first day of COVID-19 was noted to be the most likely time to encounter ocular-related manifestations (25%) as reported among those who complained of eye manifestations during the time of infection. While within the 13% who had the complaints after recovery, the time was more commonly reported to be after a month of recovery (41%) or after a week (25%). (Figure 1)

**Figure 1.**
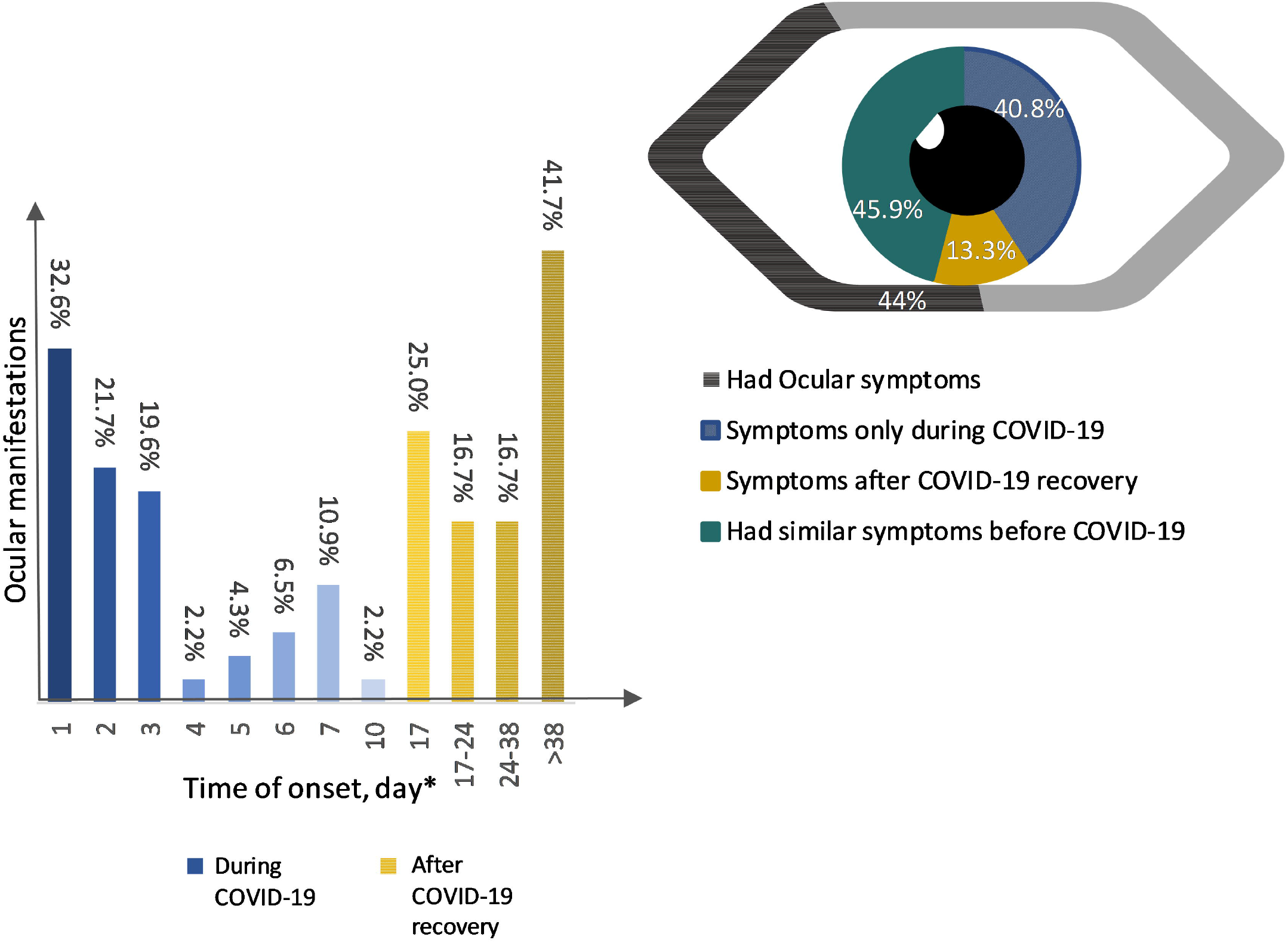
Ocular manifestations among COVID-19 participants and time of onset since COVID-19 infection diagnosis. Figure footnote: *Days since the first day of COVID-19

Non-ocular COVID-19 symptoms concerning the ocular ones were analyzed and found that these ocular manifestations were more significantly co-existed with arthralgia (OR, 95%CI; 1.77 (1.04-3.04) and chills (OR, 95%; 2.3 (1.24-4.2). As for the severity of COVID-19, no association was noted. Other related factors were longer periods of mask use (p-value=0,018), existing chronic eye diseases (p-value=0.001). (Table 3)

**Table 3.** General and COVID-19 related factors associated with ocular manifestations among COVID-19 participants. (n=276)

| Variables | With ocular<br>manifestations (n=111) | Without ocular<br>manifestation<br>(n=141) | p-value* |
| --- | --- | --- | --- |
| General characteristics: |  |  |  |
| Age, years |  |  | 0.057 |
| 18-30 | 73 (65.8%) | 71 (50.7%) |  |
| 31-45 | 22 (19.8%) | 40 (28.6%) |  |
| >45 | 16 (14.2%) | 29 (20.7%) |  |
| Gender |  |  | <0.001 |
| Female | 93 (74.4%) | 70 (53.3%) |  |
| Chronic diseases | 13 (12%) | 19 (13.5%) | 0.67 |
| Hypertension | 5 (4.5%) | 7 (5%) | 0.87 |
| Diabetes | 7 (6.3%) | 15 (10.6%) | 0.27 |
| CVD | 7 (6.3%) | 6 (4.3%) | 0.47 |
| Chronic eye diseases | 28 (25.2%) | 13 (9.2%) | 0.001 |
| Diabetic retinopathy | 0 (0%) | 3 (2%) | 0.26 |
| Refractive errors | 23 (20.7%) | 12 (8.5%) | 0.006 |
| Cataract | 1 (0.9%) | 2 (1.4%) | 0.87 |
| Contact lenses use | 8 (7.2%) | 4 (2.8%) | 0.138 |
| Smoking status |  |  | 0.76 |
| Current Smoker | 20 (18%) | 43 (30.5%) |  |
| Previous smoker | 3 (2.7%) | 3 (2.1%) |  |
| COVID-19 related: |  |  |  |
| Time of diagnosis |  |  | >0.001 |
| Currently active | 6 (6.1%) | 10 (8.8%) |  |
| During the past 3 months | 61 (61.6%) | 33 (29.6%) |  |
| During 3-6 months | 32 (32.3%) | 71 (62.3%) |  |
| previously |  |  |  |
| Disease severity |  |  | 0.43 |
| Asymptomatic | 2 (4.5%) | 2 (4.2%) |  |
| Mild | 10 (22.7%) | 18 (37.5%) |  |
| Moderate | 27 (61.4%) | 24 (50%) |  |
| Severe | 5 (11.4%) | 3 (6.3%) |  |
| Critical | 1 (1.1) | 0 |  |
| COVID-19 symptoms |  |  |  |
| Fever | 25 (17.7%) | 25 (22.5%) | 0.34 |
| Headache | 72 (64.9%) | 84 (59.6%) | 0.39 |
| Arthralgia | 43 (38.7%) | 37 (26.2%) | 0.034 |
| Myalgia | 40 (36%) | 39 (27.7%) | 0.155 |
| Dyspnea | 2 (1.8%) | 1 (0.7%) | 0.43 |
| Chills | 33 (29.7%) | 22 (15.6%) | 0.007 |
| Flu-like symptoms | 11 (9.9%) | 12 (8.5%) | 0.7 |
| Anosmia | 3 (2.7%) | 7 (5%) | 0.36 |
| Ageusia | 2 (1.8%) | 4 (2.8%) | 0.7 |
| Diarrhea | 4 (3.6%) | 2 (1.4%) | 0.4 |
| Cough | 3 (2.7%) | 0 (0%) | 0.08 |
| Other Ocular related factors: |  |  |  |
| Long office hours | 65 (58.6%) | 46 (41.4%) | 0.03 |
| health care worker | 37 (33.3%) | 29 (20.6%) | 0.03 |
| Electronic devices usage |  |  | 0.026 |
| /day | 25 (22.5%) | 28 (19.9%) |  |
| Less than 2 hours | 69 (62.2%) | 72 (50.4%) |  |
| 2-6 hours | 17 (15.3%) | 42 (29.8%) |  |
| More than 6 hours |  |  |  |
| Hours of reading /day |  |  | 0.009 |
| Less than 2 hours | 87 (78.4%) | 85 (60.3%) |  |
| 2-6 hours | 19 (17.2%) | 44 (31.2%) |  |
| More than 6 hours | 5 (4.5%) | 12 (8.5%) |  |
| Hours of mask |  |  | 0.018 |
| wearing/day | 16 (14.4%) | 14 (10.1%) |  |
| None | 35 (31.5%) | 56 (40.3%) |  |
| 1-2 | 15 (13.5%) | 31 (22.3%) |  |
| 2-4 | 20 (18%) | 25 (18%) |  |
| 4-6 | 25 (22.5%) | 13 (9.4%) |  |
| >6 |  |  |  |

CVD; cardiovascular diseases, ICU; intensive care unit

## Discussion

Ocular manifestations in this study were noted to be significantly more reported among COVID-19 infected participants during or after COVID-19 infection compared to non-COVID-19 control (44% vs 35.8%), This proposes that COVID-19 may affect the ocular tissues, supporting some hypotheses stating either that the SARS-CoV-2 virus itself can infect the eyes or secondary to the generalized immune system response to the virus.(14,17) Secondary opportunistic infection is also a likely possibility, especially known ocular pathogens.(17) The direct inoculation of the SARS-CoV-2 virus is possible due to the expression of angiotensin-converting enzyme-2 (ACE2), the main receptors for invasion of the virus to cells, which were found in aqueous humour, conjunctiva and cornea.(6,14,17)

The most frequent ocular symptoms among COVID-19 participants in this study were subjective eye dryness, burning, and itching. However, eye discharges, feeling of dryness and photosensitivity, in particular, seemed to be associated with COVID-19 compared to control. Akçay et al. assessed 1083 COVID-19 inpatients and outpatients and found the most ocular symptoms were sore eye and burning sensation (6%), with 2.6% of them had clinical conjunctivitis.(18) While in this study if used the similar definition of conjunctivitis, the incidence for COVID-19 patients was 17%, higher than Akçay et al study. As the majority of ocular symptoms in this study were in the first week, particular the first day (25%), and 10% were after recovery, as opposite to Akçay et al’ report, that began in the second week of COVID-19 in 71% of patients, and their incidences were reported to be higher among hospitalized patients, but in this study, no favour for the severity of COVID-19 was found. (18) Hong et al. study had investigated ocular symptoms on 56 patients using OSDI and Salisbury Eye Evaluation Questionnaire (SEEQ), in which they found that 27% of patients had aggravated eye irritation symptoms after COVID-19. Based on OSDI, 25% had dry eyes with a significant difference between OSDI before and after COVID-19. (17) In comparison to this study results, dry eye incidence based on OSDI was 44%, with no significance between COVID-19 and non-COVID-19. The pooled prevalence of ocular symptoms in a recent meta-analysis was 11.3%, with foreign body and dry eye being the most prevalent.(19) Hence, findings in this study were in were within the higher reported quartile, though this study was the only one to have a normal control group and including the post-COVID-19 period. Therefore, if counting incidence of eye manifestations during the time of COVID-19 and excluding after recovery, as in these studies, the incidence would be in range with these reported ones. Moreover, since 62% of those reported ocular symptoms and had COVID-19 was infected within the past 3 months, indicating the possibility of higher percentages among those more than 3 months but were not able to recall, suggesting the need for a prolonged prospective study to specifically measured this incidence.

The importance of such observation is that it may have a contribution to the early identification of COVID-19 patients, who may present with ocular findings, and seek ophthalmologists and other physicians for. Moreover, hypotheses are rising in that COVID-19 infection may transmit through tears, and that eyes may be the portal of entry, and subsequently causing direct ocular complaints and perhaps before any other as reported in previous studies.(6,20) This also emphasizes the importance of avoiding eye contact which may prevent the virus entry and ocular infection and subsequently these symptoms.(21) As shown in Chen et al. study, frequent eye contact was associated with more ocular congestion (OR, 95% CI; 4.01, (1.11–14.55)).(6)

The occurrence of ocular manifestations among COVID-19 in this study seemed to mostly accompany chills and arthralgia, with more tendency to arise appeared among females, healthcare workers, those with chronic eye diseases and contact lenses users. And long office hour workers. This result of healthcare workers being at more risk, especially ophthalmologists and any physicians with close patient contacts, highlights the need for special measurements such as eye protection for these prone groups.(21) Ocular symptoms among extended periods of mask-wearing in this study were associated with OSDI score-based eye dryness, and subjective complaints of eye discharges, forging body sensation and eye pain. Mask-associated ocular symptoms mainly dry eye, during the COVID-19 pandemic, was previously explored in a large survey in Italy and found that mask-associated dry eye (MADE) was 18.3%, of which 26.1% had increasing eye symptoms after wearing face masks.(15) This complies with this study results, though the prevalence of dry eye among extended face mask use was 60.8% and not regarded to whether symptoms worsen with face masks, but were compared in terms of hours of use. Moreover, in their study, the COVID-19 status of participants was not explored or correlated with face-mask, which showed to confound some results. Accordingly, this study findings of mask-associated dryness and irritation especially in face mask users above 6 hours, alert the general public and care providers, when need to wear masks for prolonged periods, for the need to be more cautious and some methods like using lubricant and wearing eye protection.

This study was unique for the combination between the direct COVID-19 effect and the effect of face mask use due to the COVID-19 pandemic while investigating the association with ocular manifestations. As the exact relation and the underlying image won’t be clear unless gathered all together. Moreover, this was the first study that had a healthy control group, in order to have more reasonable comparisons that can reflect the general population. However, the study had many limitations, as it was based on a questionnaire, all results were subjective, and findings were not confirmed clinically, like by slit-lamp examination or Schirmer’s test. Additionally, its retrospective nature made it prone to recall bias, especially underestimating the ocular complaints incidence for considering eye manifestations are trifles, compared to other COVID-19 symptoms, though six months were chosen as recall memory would still be optimal. the control group had no tests as well, and so the possibility of someone having an asymptomatic COVID-19 was possible. But in both arms, symptoms of COVID-19 were investigated and the dilemmatic COVID-19 state cases were excluded.

In conclusion, the COVID-19 pandemic had sequences that could even involve the eyes, causing irritations from the virus itself, and more broadly was responsible for the evolving of mask-associated dry eye, as part of its preventive measures. All of these should not be ignored and be considered when dealing with patients complaining of eye manifestations, and when need to wear masks for extended periods, lubricant or other methods would be suggested to avoid mask associated dryness.

## Data Availability

Data are available upon request from the corresponding author.

## Data availability statement

Data are available upon request from the corresponding author.

## Ethics statement

Consent was obtained from participants, within the questionnaire after describing the study and its objectives and ensuring the participants’ data privacy. The study gained ethical approval that was conducted in accordance with the Declaration of Helsinki by AL-Najah National University’ Ethics Committee.

